# SARS-CoV-2 breakthrough infection during the Delta-dominant epidemic and neutralizing antibodies against Omicron in comparison with the third dose of BNT162b2: a matched analysis

**DOI:** 10.1101/2022.06.21.22276682

**Authors:** Shohei Yamamoto, Kouki Matsuda, Kenji Maeda, Yusuke Oshiro, Natsumi Inamura, Tetsuya Mizoue, Maki Konishi, Junko S. Takeuchi, Kumi Horii, Mitsuru Ozeki, Haruhito Sugiyama, Hiroaki Mitsuya, Wataru Sugiura, Norio Ohmagari

## Abstract

**Background:** Longitudinal data are lacking to compare booster effects of Delta breakthrough infection versus the third vaccine dose on neutralizing antibodies (NAb) against Omicron.

**Methods:** Participants were the staff of a national research and medical institution in Tokyo who attended serological surveys on June 2021 (baseline) and December 2021 (follow-up); in between, the Delta-dominant epidemic occurred. Of 844 participants who were infection-naïve and had received two doses of BNT162b2 at baseline, we identified 11 breakthrough infections during the follow-up. One control matched to each case was randomly selected from those who completed the booster vaccine and those who were unboosted by the follow-up. We used the generalized estimating equation model to compare live-virus NAb against Wuhan, Delta, and Omicron across groups.

**Results:** Persons who experienced breakthrough infection showed marked increases in NAb titers against Wuhan (4.1-fold) and Delta (5.5-fold), and 64% had detectable NAb against Omicron at follow-up, although the NAb against Omicron after breakthrough infection was 6.7- and 5.2-fold lower than that against Wuhan and Delta, respectively. The increase was apparent only in symptomatic cases and as high as in the third vaccine recipients. In contrast, these titers largely decreased (Wuhan, Delta) or remained undetected (Omicron) at follow-up in infection-naïve and unboosted persons.

**Conclusions:** Symptomatic breakthrough infection during the Delta predominant wave was associated with significant increases in NAb against Wuhan, Delta, and Omicron, similar to the third BNT162b2 vaccine. Given the much lower cross-NAb against Omicron than other virus types, however, infection prevention measures must be continued irrespective of vaccine and infection history while the immune evasive variants are circulating.

**Key points:** Symptomatic, not asymptomatic, SARS-CoV-2 breakthrough infection after the second BNT162b2 vaccination during the Delta-predominant wave enhanced neutralizing antibodies against Wuhan, Delta, and Omicron comparable to the three vaccine doses, although immunity against Omicron was much lower than Wuhan and Delta.

## Introduction

Clinical trials showed that the mRNA-based vaccine was highly effective in reducing the risk of morbidity and mortality of coronavirus disease 2019 (COVID-19) [1]. The waning of vaccine-induced immunogenicity over time [2, 3] and the emergence of variants of concern (VOCs) with a high potential for immune evasion [4], however, have led to a marked increase in breakthrough infections and urged many countries to adopt the booster (third) vaccine campaign. As a result, the source of immunity against SARS-CoV-2 became diverse among people through their history of vaccination and infection. In this situation, there would be a need for quantitative data on immunogenicity according to infection/vaccination histories, which would help their decision on whether to receive a booster dose to prevent forthcoming variants.

A few studies showed that among infection-naïve two-dose vaccine recipients, neutralizing antibody (NAb) levels against VOCs, including the Omicron variant, were increased in persons who experienced Delta breakthrough infection or had received the third vaccine dose [5-7]. These studies, however, have some methodological limitations. No study considered potential modifiers of vaccine response such as age and sex in selecting controls or analyzing data and assessed the change of NAb between pre- and post-breakthrough infection. It thus remains elusive to what extent the NAb increased after breakthrough infection relative to the baseline. Additionally, few studies [6] analyzed the data by COVID-19 symptom, despite consistent evidence that higher antibody concentrations among patients with symptomatic infection than those with asymptomatic infection [8].

The objective of the present study was thus to assess live-virus NAb against the Wuhan, Delta, and Omicron variants among two-dose BNT162b2 vaccine recipients who experienced breakthrough infection during the Delta variant predominant wave, compared with unboosted and boosted infection-naïve individuals in a well-defined cohort of health care workers while addressing the issues mentioned above.

## Methods

### Study setting and case-control selection

We used data and samples from a repeated serological study among the staff of the National Center for Global Health and Medicine, Japan (NCGM). The detail of the study has been described elsewhere [9]. Written informed consent was obtained from all participants. The study procedure was approved by the NCGM Ethics Committee (approval number: NCGM-G-003598).

Of 948 participants who attended both baseline (June 2021) and follow-up (December 2021) surveys (**Figure 1**), 844 had received two doses of BNT162b2 vaccine and were infection-naïve (i.e., no history of COVID-19 and negative with anti-SARS-CoV-2 nucleocapsid protein assays [both Abbott and Roche assays]) at baseline. Of these, we excluded 8 participants who received a vaccine other than BNT162b2 as the booster, leaving 836 included in the source population (528 had received two doses and 308 had received three doses by follow-up). Of those, we identified 11 patients who had the breakthrough infection during follow-up (7 cases were ascertained via in-house COVID-19 registry, and 4 cases were additionally identified with antibody test) and attended the follow-up survey without receiving the booster dose. All cases were assumed to be infected with the Delta variant, given its predominant circulation during the study period in Japan. For each case, we randomly selected infection-naïve controls from those who did not receive the third dose of the BNT162b2 and those who received the third dose during the study period, while individually matching on sex, age (± 3 years), spike IgG antibody titer (± 10%), and the interval between the second vaccination and blood sampling at baseline.

**Figure 1.**
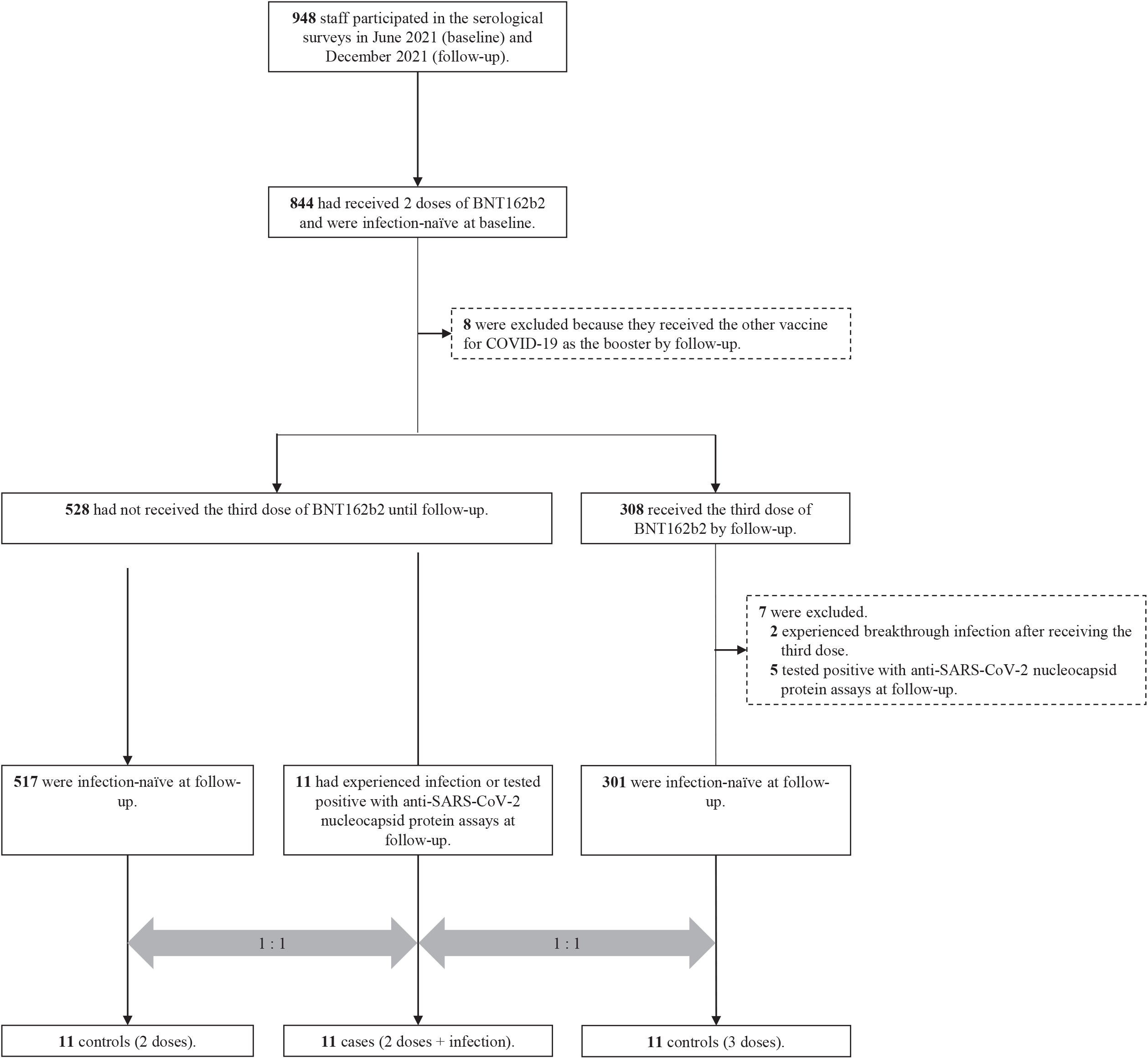
Flowchart for the case-control matching. Infection-naïve were defined as having no history of COVID-19 and being negative with anti-SARS-CoV-2 nucleocapsid protein assays (both Abbott and Roche assays). Abbreviations: COVID-19, coronavirus disease 2019; SARS-CoV-2, severe acute respiratory syndrome coronavirus 2.

### Neutralizing antibody testing

The NAb in serum was determined by quantifying the serum-mediated suppression of the cytopathic effect (CPE) of each SARS-CoV-2 strain in VeroE6_TMPRSS2_ cells [10, 11]. The obtained routes of the cells and each virus are described in **Supplemental Text 1**. Each serum sample was 4-fold serially diluted in a culture medium. The diluted sera were incubated with 50% tissue culture infectious dose (TCID_50_) of the virus at 37°C for 20 minutes (final serum dilution range of 1:40 to 1:25000), after which the serum-virus mixtures were inoculated with VeroE6_TMPRSS2_ cells (1.0×10^4^/well) in 96-well plates. The SARS-CoV-2 strains used in these assays are as follows: a Wuhan, wild-type strain (SARS-CoV-2^05-2N^) [12], a Delta variant (SARS-CoV-2^TKYTK1734/2021^), and an Omicron BA.1 variant (SARS-CoV-2^TKYX00012/2021^). After culturing the cells for 3 to 5 days, the levels of CPE observed in SARS-CoV-2–exposed cells were determined using the WST-8 assay using the Cell Counting Kit-8 (Dojindo, Kumamoto, Japan). The serum dilution that gave 50% inhibition of CPE was defined as the 50% neutralization titer (NT_50_). Each serum sample was tested in duplicate, and the average value was used for analysis.

### SARS-CoV-2 antibody testing

We assessed anti–SARS-CoV-2 antibodies for all participants at baseline and follow-up and retrieved those data for the case-control subsets. We quantitatively measured antibodies against the receptor-binding domain (RBD) of the SARS-CoV-2 spike protein by using the AdviseDx SARS-CoV-2 IgG II assay (Abbott) (immunoglobulin [Ig] G [IgG]) and Elecsys^®^ Anti-SARS-CoV-2 S RUO (Roche) (including IgG).

We also qualitatively measured antibodies against SARS-CoV-2 nucleocapsid protein using the SARS-CoV-2 IgG assay (Abbott) and Elecsys^®^ Anti-SARS-CoV-2 RUO (Roche), and used these data to exclude those with the possible infection before the baseline survey and to identify those who experienced breakthrough infection during the follow-up period. The sensitivity and specificity were 100% and 99.9%, respectively, for the Abbott assay [13], and 99.5% and 99.8%, respectively, for the Roche assay [14].

### Statistical analysis

We compared the characteristics between patients with breakthrough infection and their matched controls using the Kruskal-Wallis test or Fisher’s exact test. To examine the group difference of the changes in neutralizing and spike antibody levels from baseline to follow-up, we used a generalized estimating equation (GEE) with unstructured correlation structures and the robust variance estimator. Neutralizing and anti-spike antibody titers were log-transformed before analysis. Independent variables included were the matching indicator, time (baseline or follow-up), and interaction terms between group and time. The estimated effects of covariates were back-transformed and presented as ratios of geometric means. We repeated the GEE model to compare the neutralizing and spike antibody titers among the following three groups: those who had symptomatic breakthrough infections, asymptomatic breakthrough infections, or received three vaccinations. We also run the GEE model with independent correlation structures to compare NAb titers against the Wuhan, Delta, and Omicron BA.1 within each of the following four groups: a group of all participants at baseline (n=33) and three groups at follow-up (those who received only two doses (n=11), those who experienced breakthrough infection (n=11), and those who received three doses (n=11)).

For analyses, values below or above the limit of detection (LOD) for NAb titers (NT_50_<40) and spike antibody titers with Roche assay (titer >25000 U/mL) were given the LOD value, respectively. Statistical analysis was performed using Stata version 17.0 (StataCorp LLC), and graphics were made by GraphPad Prism 9 (GraphPad, Inc). All P values were 2-sided, and P < 0.05 was considered statistically significant.

## Results

### Characteristics of breakthrough infection cases

Of the 11 patients of breakthrough infection, 55% were male, the median age was 26 (interquartile range [IQR]: 24–29) years, and the median body mass index was 21 (IQR: 20–22) kg/m^2^. Their major occupations were nurses (55%), allied healthcare professionals (18%), and doctors (5%), and 45% were engaged in COVID-19-related work. The median intervals from the second BNT162b2 vaccine to baseline and follow-up survey were 63 (IQR: 39-71) and 247 (IQR: 218-250) days, respectively. At baseline, the median spike antibody titers measured with Abbott and Roche assays were 3739 (IQR: 2435–9555) AU/mL and 935 (IQR: 762–1260) U/mL, respectively. The median NAb titers against Wuhan and Delta were 156 (IQR: 40-347) and 101 (IQR: 42-162) NT_50_, respectively, while no participant had detectable NAb against Omicron BA.1. These figures were similar across groups (**Table 1**).

**Table 1.**
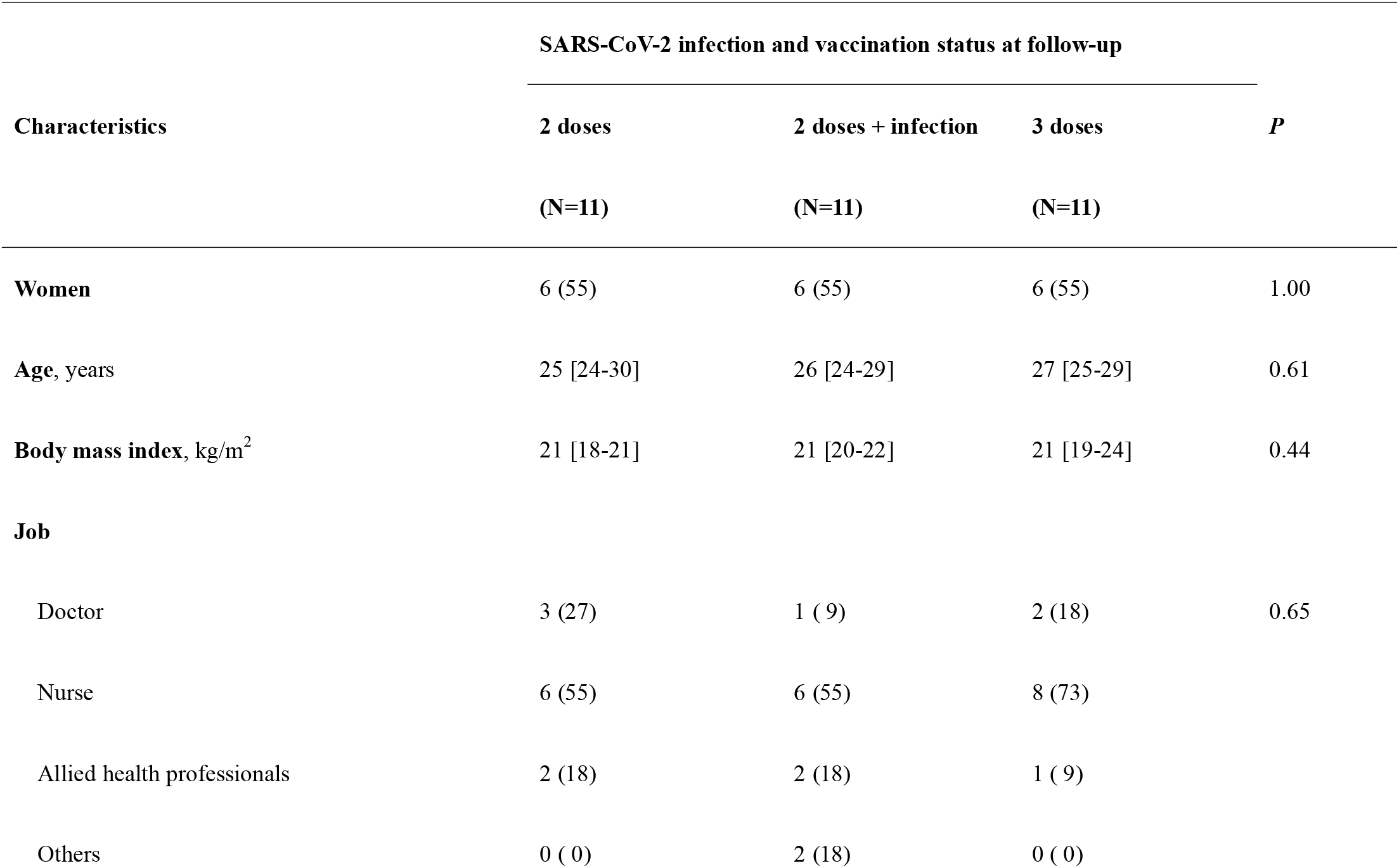
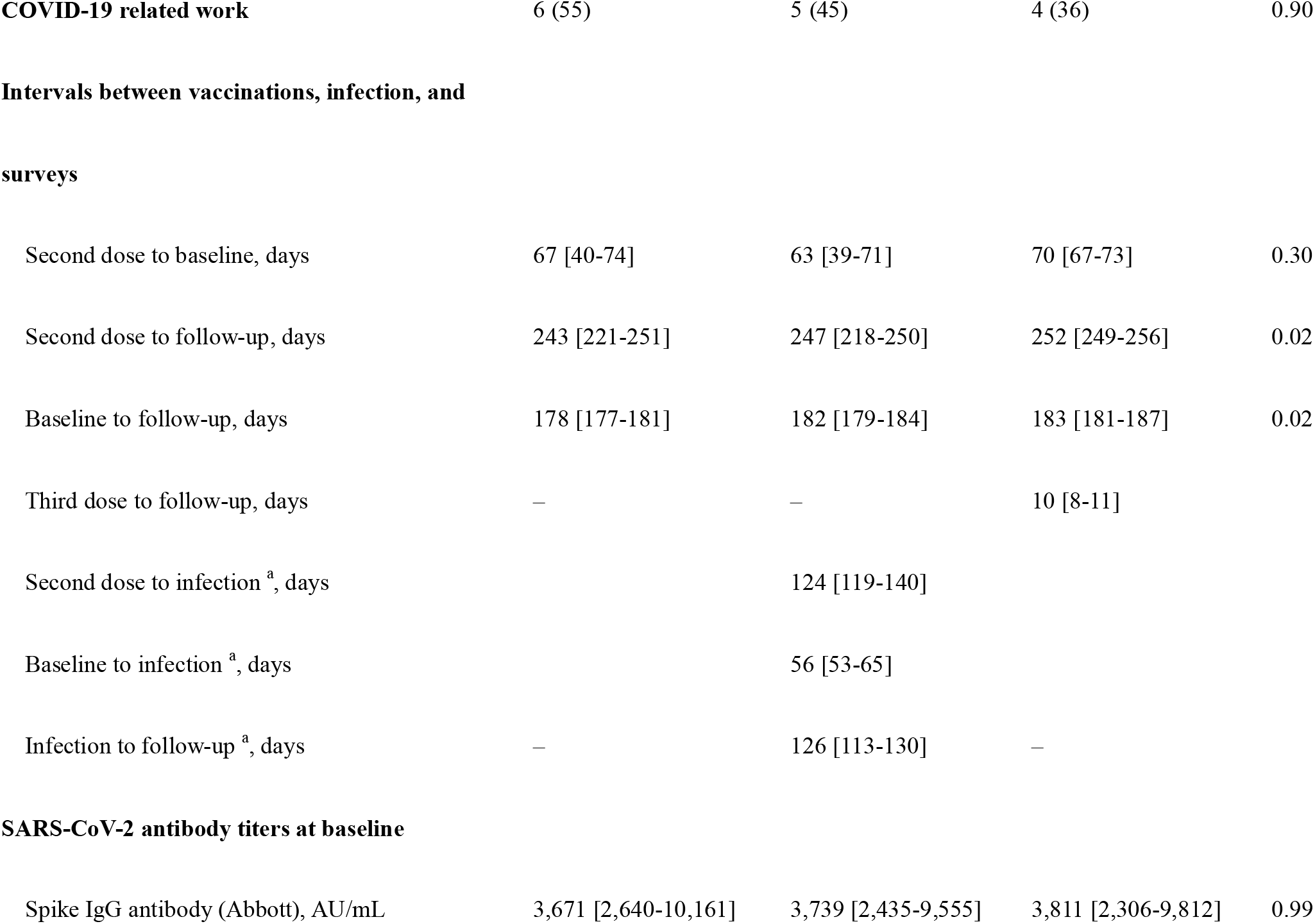
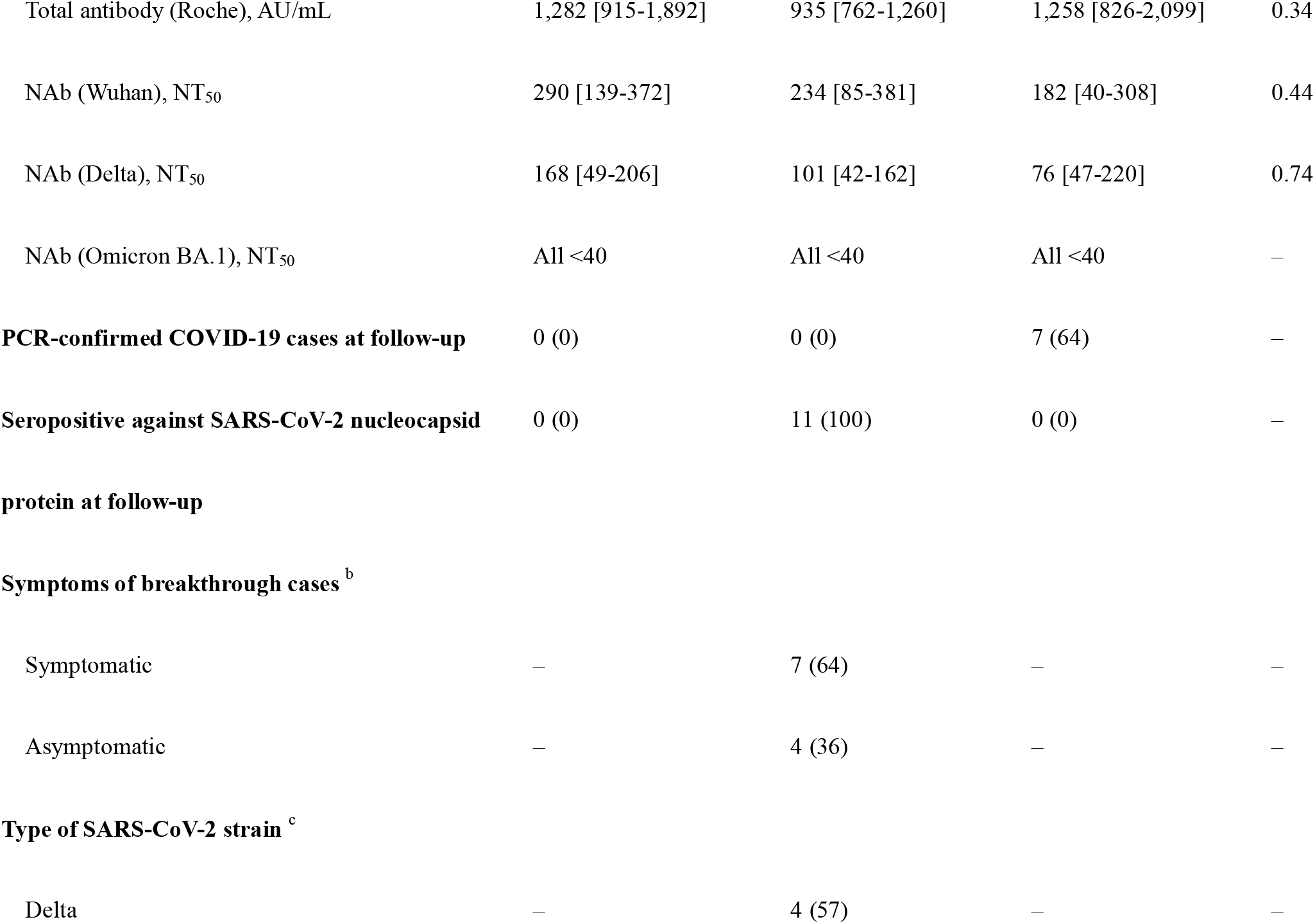
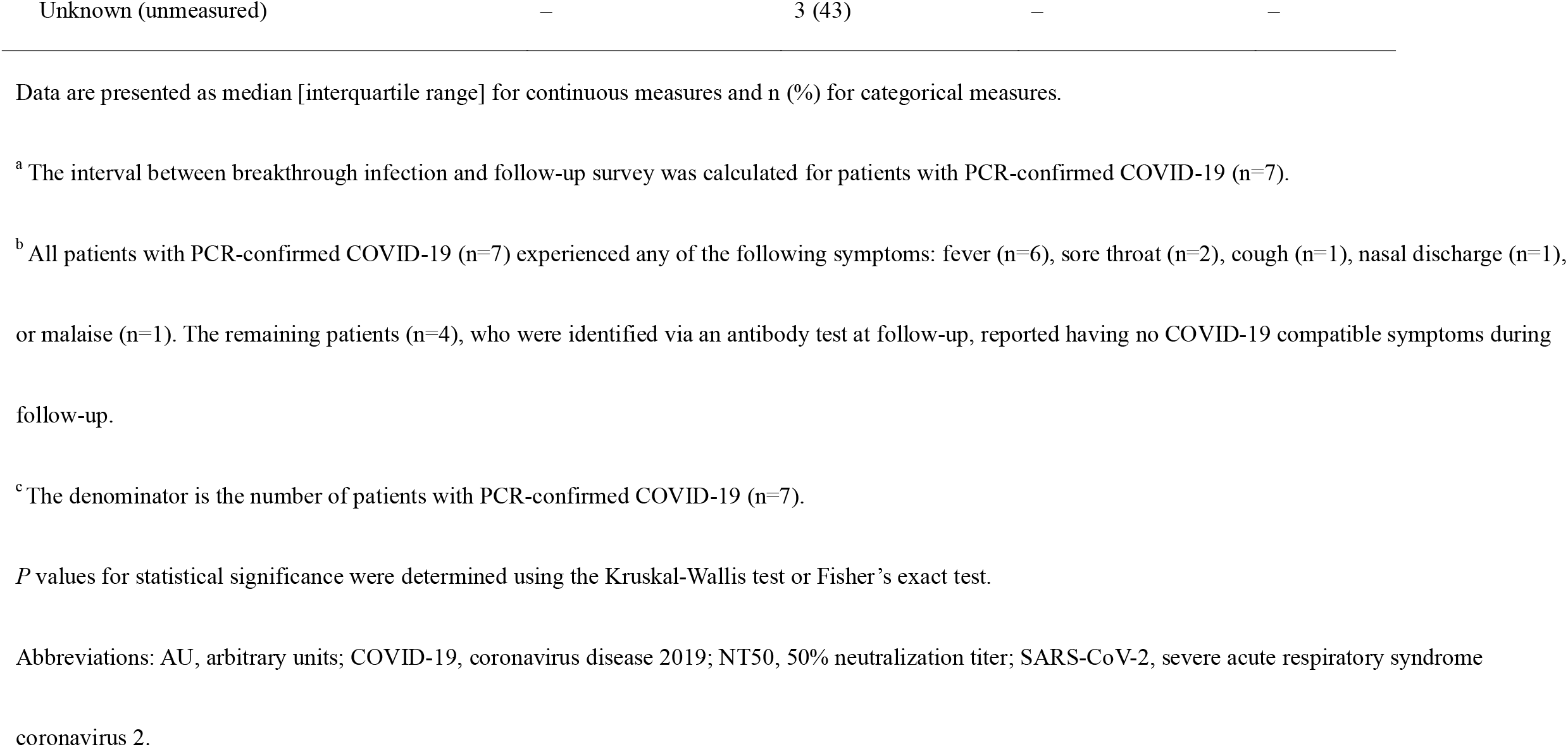
Characteristics of the study participants

| Characteristics | SARS-CoV-2 infection and vaccination status at follow-up |  |  | *P* |
| --- | --- | --- | --- | --- |
|  | 2 doses | 2 doses + infection | 3 doses |  |
|  | (N=11) | (N=11) | (N=11) |  |
| **Women** | 6 (55) | 6 (55) | 6 (55) | 1.00 |
| **Age, years** | 25 [24-30] | 26 [24-29] | 27 [25-29] | 0.61 |
| **Body mass index, kg/m<sup>2</sup>** | 21 [18-21] | 21 [20-22] | 21 [19-24] | 0.44 |
| **Job** |  |  |  |  |
| Doctor | 3 (27) | 1 ( 9) | 2 (18) | 0.65 |
| Nurse | 6 (55) | 6 (55) | 8 (73) |  |
| Allied health professionals | 2 (18) | 2 (18) | 1 ( 9) |  |
| Others | 0 ( 0) | 2 (18) | 0 ( 0) |  |
| **COVID-19 related work** | 6 (55) | 5 (45) | 4 (36) | 0.90 |
| **Intervals between vaccinations, infection, and surveys** |  |  |  |  |
| Second dose to baseline, days | 67 [40-74] | 63 [39-71] | 70 [67-73] | 0.30 |
| Second dose to follow-up, days | 243 [221-251] | 247 [218-250] | 252 [249-256] | 0.02 |
| Baseline to follow-up, days | 178 [177-181] | 182 [179-184] | 183 [181-187] | 0.02 |
| Third dose to follow-up, days | — | — | 10 [8-11] |  |
| Second dose to infection <sup>a</sup>, days |  | 124 [119-140] |  |  |
| Baseline to infection <sup>a</sup>, days |  | 56 [53-65] |  |  |
| Infection to follow-up <sup>a</sup>, days | — | 126 [113-130] | — |  |
| **SARS-CoV-2 antibody titers at baseline** |  |  |  |  |
| Spike IgG antibody (Abbott), AU/mL | 3,671 [2,640-10,161] | 3,739 [2,435-9,555] | 3,811 [2,306-9,812] | 0.99 |
| Total antibody (Roche), AU/mL | 1,282 [915-1,892] | 935 [762-1,260] | 1,258 [826-2,099] | 0.34 |
| NAb (Wuhan), NT<sub>50</sub> | 290 [139-372] | 234 [85-381] | 182 [40-308] | 0.44 |
| NAb (Delta), NT<sub>50</sub> | 168 [49-206] | 101 [42-162] | 76 [47-220] | 0.74 |
| NAb (Omicron BA.1), NT<sub>50</sub> | All <40 | All <40 | All <40 | – |
| **PCR-confirmed COVID-19 cases at follow-up** | 0 (0) | 0 (0) | 7 (64) | – |
| **Seropositive against SARS-CoV-2 nucleocapsid protein at follow-up** | 0 (0) | 11 (100) | 0 (0) | – |
| **Symptoms of breakthrough cases <sup>b</sup>** |  |  |  |  |
| Symptomatic | – | 7 (64) | – | – |
| Asymptomatic | – | 4 (36) | – | – |
| **Type of SARS-CoV-2 strain <sup>c</sup>** |  |  |  |  |
| Delta | – | 4 (57) | – | – |
| Unknown (unmeasured) | – | 3 (43) | – | – |

Data are presented as median [interquartile range] for continuous measures and n (%) for categorical measures.
<sup>a</sup> The interval between breakthrough infection and follow-up survey was calculated for patients with PCR-confirmed COVID-19 (n=7).
<sup>b</sup> All patients with PCR-confirmed COVID-19 (n=7) experienced any of the following symptoms: fever (n=6), sore throat (n=2), cough (n=1), nasal discharge (n=1), or malaise (n=1). The remaining patients (n=4), who were identified via an antibody test at follow-up, reported having no COVID-19 compatible symptoms during follow-up.
<sup>c</sup> The denominator is the number of patients with PCR-confirmed COVID-19 (n=7).
*P* values for statistical significance were determined using the Kruskal-Wallis test or Fisher's exact test.
Abbreviations: AU, arbitrary units; COVID-19, coronavirus disease 2019; NT50, 50% neutralization titer; SARS-CoV-2, severe acute respiratory syndrome coronavirus 2.

For 7 persons with a history of PCR-confirmed breakthrough infection, the median interval between the second dose and infection and between infection and follow-up survey was 124 (IQR: 119–140) and 126 (113-130) days, respectively. All these patients experienced any of the following symptoms: fever (n=6), sore throat (n=2), cough (n=1), nasal discharge (n=1), or malaise (n=1), and were seropositive with anti-SARS-CoV-2 nucleocapsid protein assays at the follow-up survey. Data on virus strain were available for 4 of 7 (57%) cases (all were the Delta variant). The remaining 4 cases were identified via anti-SARS-CoV-2 N protein assays at the follow-up survey; all reported having no COVID-19 compatible symptoms during follow-up.

### Antibody titers waned over time and were boosted by breakthrough infection and the third dose

During the follow-up, NAb titers against Wuhan showed a 4.1-fold and 10.9-fold increase in those who experienced breakthrough infection and those who received the third dose, respectively. In contrast, it became undetectable in all unboosted infection-naïve individuals (**Figure 2A**). At the follow-up survey, persons with breakthrough infections had a 2.2-fold lower, albeit statistically not significant, geometric mean titers (GMT) against Wuhan (723, 95% CI: 261–2001) than the third-dose vaccine recipients (1580, 95% CI: 1043–2395). The NAb titer against Delta was 2.1-fold decreased in those who received only two doses, whereas it showed 5.5-fold and 13.8-fold increases in those with breakthrough infection and those with three doses, respectively (**Figure 2B**). Consequently, persons with breakthrough infection had a 2.8-fold lower, albeit statistically not significant, GMT against Delta (GMT: 564, 95% CI: 187–1703) than those who received the third dose (GMT: 1563, 95% CI: 873–2801) at the follow-up survey. The NAb titer against Omicron BA.1, which was not detectable in all groups at baseline, was quantifiable in two-thirds of those who experienced breakthrough infection (GMT: 108, 95% CI: 72–163) and all persons who received the third dose (GMT: 152, 95% CI: 101–228) (**Figure 2C**).

**Figure 2.**
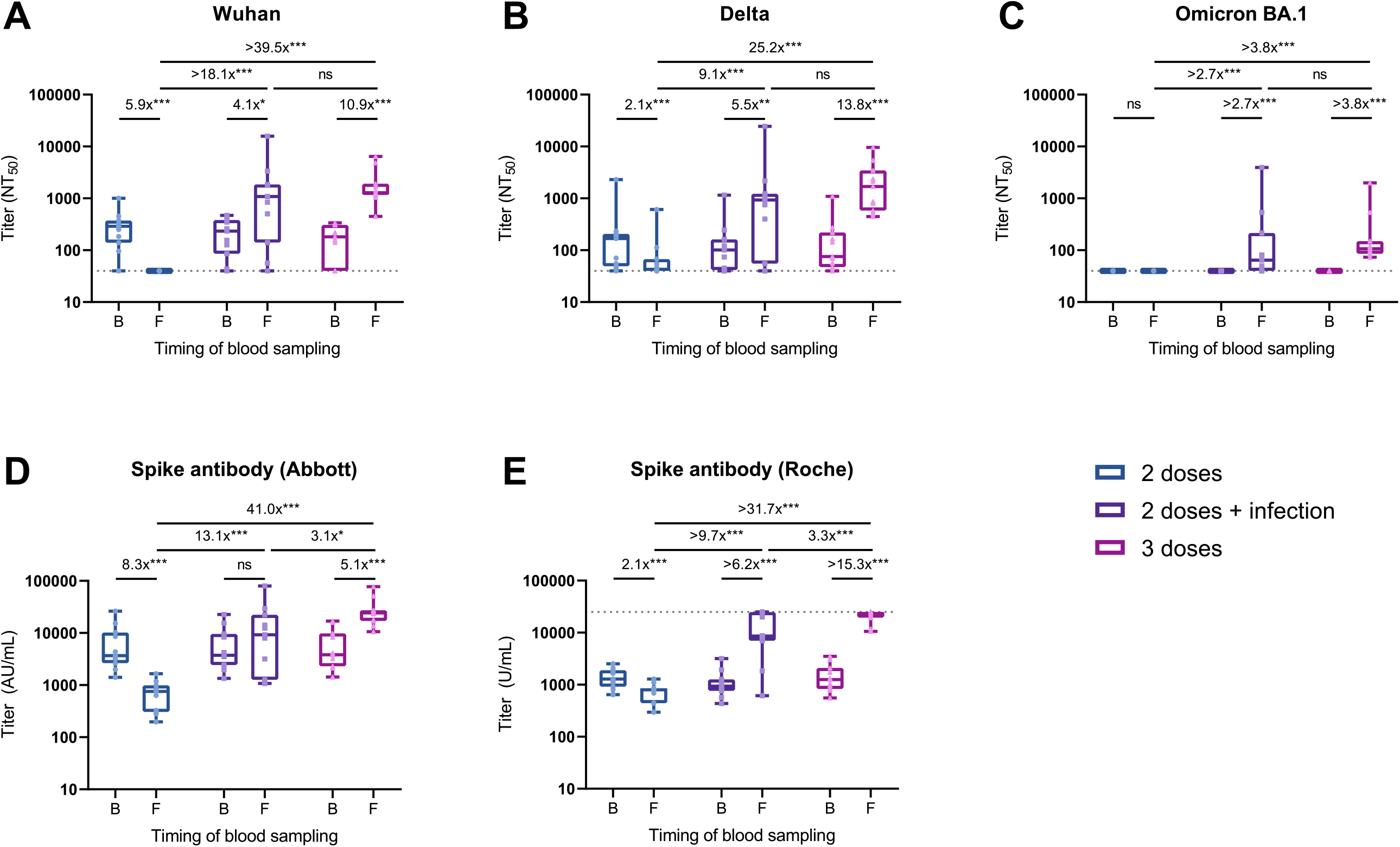
Change in the neutralizing and spike antibody titers in individuals who experienced breakthrough infection, received the booster vaccine, or were unboosted during follow-up. Shown are NAb titers against the original Wuhan strain (**A**), the Delta variant (**B**), and the Omicron BA.1 variant (**C**) determined by 50% focus reduction neutralization test (FRNT_50_) using the serum at baseline and follow-up. Also shown are anti-spike antibody titers measured with the Abbott reagent (**D**) and the Roche reagent (**E**) at baseline and follow-up. Box plots show the median, interquartile range, and full range. The dushed horizontal lines indicate the LOD in the present analysis (NT_50_<40 in FRNT_50_ and U/mL>25000 in Roche assay). The fold-change values are estimated ratios of geometric means for antibody titers based on the GEE model (ns: not significant; *P<0.05; **P<0.01; ***P<0.001). Abbreviations: AU, arbitrary units; B, baseline; F, follow-up; GEE, generalized estimating equation; LOD, limits of detection; NT_50_, 50% neutralization titer; SARS-CoV-2, severe acute respiratory syndrome coronavirus 2.

Mean spike antibody titer with Abbott assay showed an 8.3-fold decrease in those who did not receive the booster dose, no change in those with breakthrough infections, and a 5.1-fold increase in the third vaccine recipients (**Figure 2 D**). As a consequence, persons with breakthrough infection had a 3.1-fold lower mean titer than those who received the third dose at follow-up. Mean spike antibody titer with Roche assay showed a 2.1-fold decrease in those who did not receive the booster dose, whereas it showed at least 6.2-fold and 15.3-fold increase in those who experienced breakthrough infection and the recipients of the third dose, respectively (**Figure 2 E**). At follow-up, persons with breakthrough infection had a 3.3-fold lower mean titer than the third-dose vaccine recipients.

### Symptomatic, but not asymptomatic, breakthrough infection cases had NAb titers comparable to the third dose recipients

All types of NAb titer were substantially increased in persons with symptomatic breakthrough infection (3.8 to 17.7 folds), whereas the titer against Wuhan showed a 3.1-fold decrease and those against Delta and Omicron BA.1 did not increase in those with asymptomatic breakthrough infection (**Figure S1**). At follow-up, persons with a history of symptomatic infection had GMT against Wuhan of 2093, Delta of 1542, and Omicron BA.1 of 151, comparable to those of the third vaccine recipients (1580, 1563, and 152 in sequential order; **Figure 3**).

**Figure 3.**
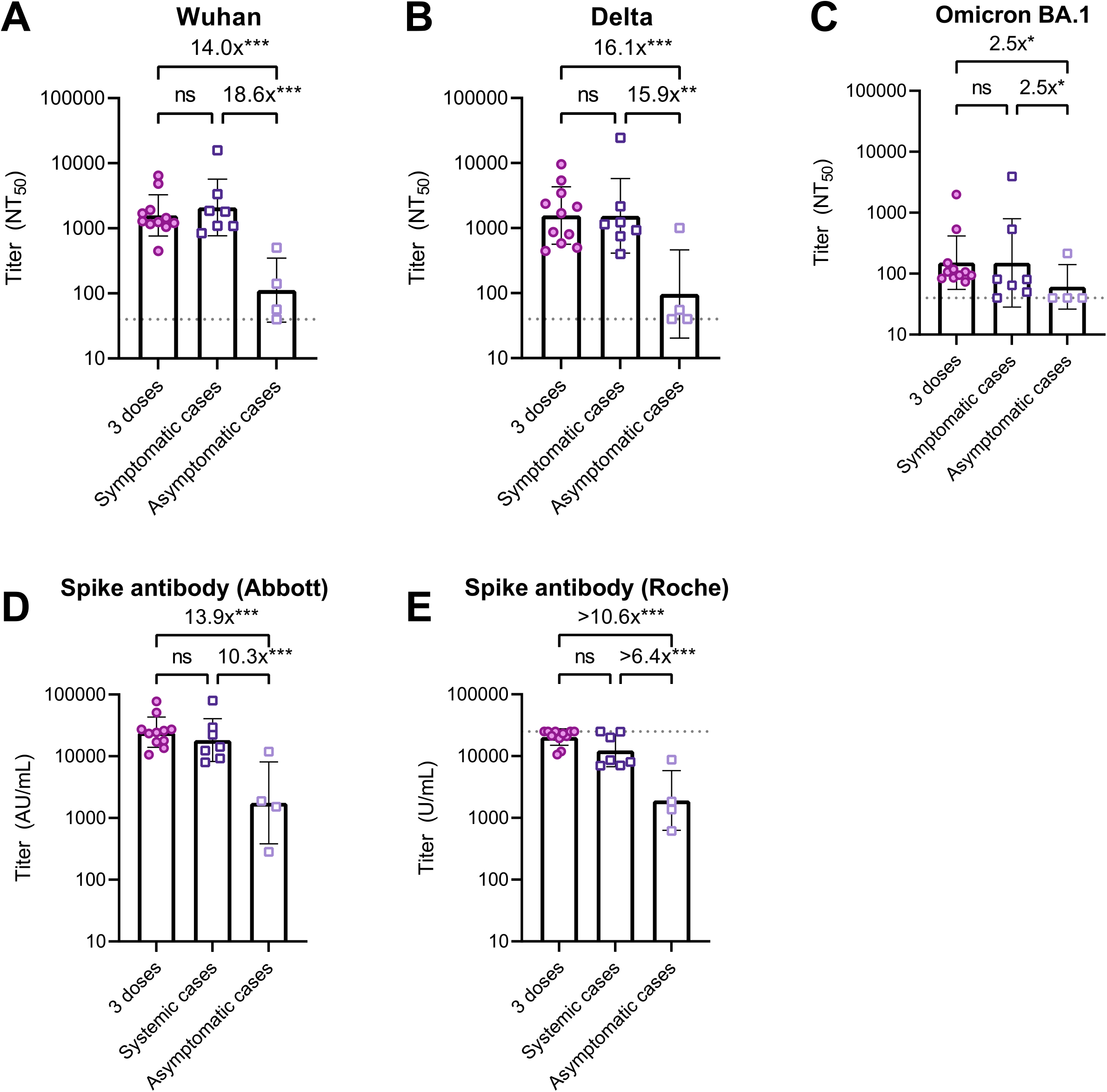
Neutralizing and spike antibody titers after three vaccine doses, symptomatic and asymptomatic breakthrough infections. Shown are NAb titers against the original Wuhan strain (**A**), the Delta variant (**B**), and the Omicron BA.1 variant (**C**) determined by 50% focus reduction neutralization test (FRNT_50_) using the serum at follow-up. Also shown are anti-spike antibody titers measured with the Abbott reagent (**D**) and the Roche reagent (**E**) using the serum at follow-up. Patients with PCR-confirmed infection were all symptomatic (n=7), while those with seropositive on any of anti-SARS-CoV-2 nucleocapsid protein assays (Abbott or Roche assays) at follow-up were all asymptomatic (n=4). The bars indicate geometric mean titers, and I-shaped bars indicate its geometric standard deviations. The dushed horizontal lines indicate the LOD for FRNT_50_ (NT_50_<40) and Roche assay (U/mL>25000) in the present analysis. Statistical significance was determined by Kruskal-Wallis and Dunn’s multiple comparison test (ns: not significant; *P<0.05; **P<0.01; ***P<0.001). Abbreviations: AU, arbitrary units; COVID-19, coronavirus disease 2019; LOD, limits of detection; NT50, 50% neutralization titer; SARS-CoV-2, severe acute respiratory syndrome coronavirus 2.

### Neutralizing capacity against Omicron BA.1 was markedly lower than those against Wuhan and Delta after breakthrough infection or third dose of vaccine

At baseline (all serum samples combined), most had a quantifiable NAb against Wuhan (85%) and Delta (89%); the geometric mean titer against Delta was 1.6-fold lower than that against Wuhan. None had a quantifiable NAb titer against Omicron BA.1 (**Figure 4**). At follow-up, NAb titer against Omicron BA.1 was markedly lower than that against Wuhan and Delta in persons who experienced the breakthrough infection (6.7- and 5.2-fold, respectively) and those who completed the third dose (10.4- and 10.3-fold, respectively) (**Figure 4**).

**Figure 4.**
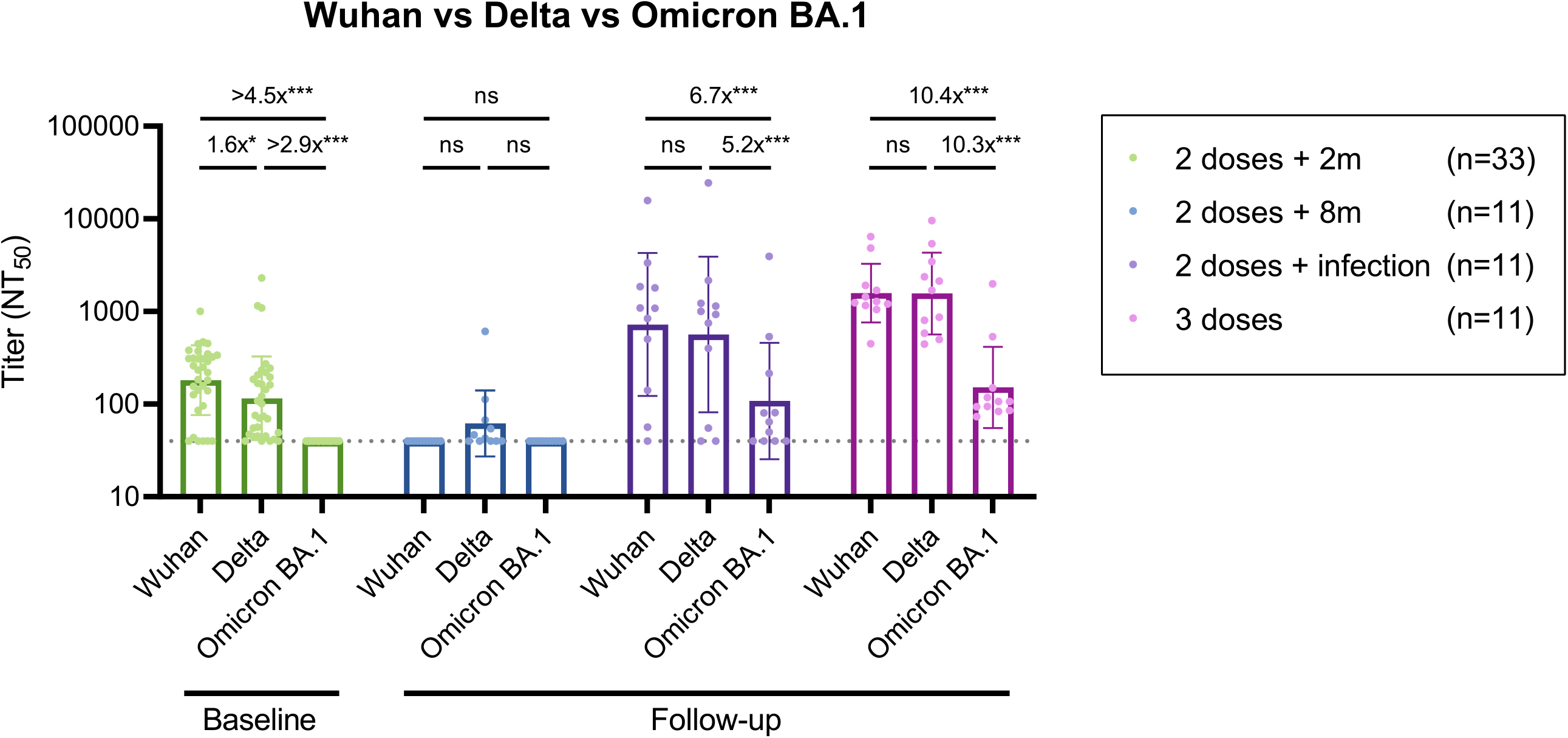
Comparison of neutralizing antibody titers against Wuhan, Delta, and Omicron BA.1 strains. Shown are the NAb titers against the original Wuhan strain, the Delta variant, and the Omicron BA.1 variant as determined by the 50% focus reduction neutralization test (FRNT_50_). The bars indicate geometric mean titers, and I-shaped bars indicate its geometric standard deviations. The dushed horizontal lines indicate the LOD (NT_50_ titer <40). Statistical significance was determined by the GEE model (ns: not significant; *P<0.05; **P<0.01; ***P<0.001). Abbreviations: GEE, generalized estimating equation; LOD, limits of detection; NT_50_, m: months, 50% neutralization titer; 95% CI, 95% confidence interval.

## Discussion

In this matched longitudinal study among infection-naïve individuals who received two doses of BNT162b2, the live-virus NAb titers against Wuhan, Delta, and Omicron BA.1 were increased among persons who were infected during the Delta-dominant epidemic period. The increase of NAb was prominent in persons with symptomatic, but not asymptomatic, breakthrough infection and was comparable to that in the third-dose vaccine recipients.

Our finding of an increase in cross-reactive NAb against Wuhan and Omicron after breakthrough infection during the Delta wave is consistent with previous reports [5-7]. With the rigorous matching of background factors, the use of paired serum samples, and the assessment of NAb against live viruses, the present study adds to confirmatory evidence in the literature regarding the role of Delta breakthrough infection in immunological cross-reactivity to Omicron. Nevertheless, we should note that their NAb titers against Omicron were much lower than those against Wuhan and Delta, probably due to frequent spike mutations in the Omicron variant [15]. This result is compatible with observational data showing frequent reinfections with Omicron among those with a history of infection with former SARS-CoV-2 strains, including Delta [16], and lowered effectiveness of three BNT162b2 doses against Omicron infection [17, 18].

The severity of COVID-19 has been suggested to correlate with post-infection immunogenicity [8]. In our analysis, NAb titers against Wuhan and Delta were increased in persons with COVID-19 compatible symptoms at breakthrough infection but not in those with asymptomatic infection (detected with antibody test only). This result is in line with a study among patients who recovered from Delta infection [6], reporting higher NAb titers against Wuhan in those with moderate to severe symptoms than in those with no or mild symptoms. More importantly, we found that among patients who were infected during the high circulation of Delta, the proportion of having a detectable NAb titer against Omicron was much higher among persons who had any symptom than those who had no symptom (86% vs. 25%), suggesting that cross-reactivity induced by Delta infection against the Omicron variant also depends on the symptom.

The limitations of this study should be acknowledged. First, the interval from SARS-CoV-2 exposure (infection or vaccination) to blood sampling markedly differed between patients with PCR-confirmed breakthrough infection (median 126 [IQR: 113–130] days) and the third vaccine recipients (median 10 [IQR: 8–11] days). Accordingly, antibody titers might be still on the rise in the former, while they may reflect waning over time in the latter. Second, viral sequence data were available for only 4 (57%) patients among PCR-confirmed COVID-19 cases. Nevertheless, we can reasonably assume that the remaining breakthrough infections are also due to the Delta variant, which accounted for more than 90% of sequenced COVID-19 samples in Japan during the follow-up (June to December 2021) [19]. Third, we did not assess cellular immune response, another important mechanism for preventing severe COVID-19 [20]. Finally, study participants were predominantly lean Japanese (median [IQR] body mass index of 21 [19–22] kg/m^2^). Caution should be exercised in generalizing the present findings to the populations with different backgrounds.

In summary, recipients of two doses of BNT162b2 who subsequently experienced symptomatic breakthrough infection (possibly with Delta) showed a substantial increase in NAb titers against Wuhan, Delta, and, to a lesser extent, Omicron BA.1, similar to those who received the third vaccine dose. The results may help those who experienced breakthrough infection in their decision making whether to receive the booster vaccine. Given their markedly lower cross-reactive NAb titers against Omicron than other virus types, however, infection prevention measures must be continued, irrespective of SARS-CoV-2 infection or vaccination history.

## Data Availability

The datasets generated and/or analyzed during the current study are not publicly available due to ethical restrictions and participant confidentiality concerns, but de-identified data are available from the corresponding author to qualified researchers on reasonable request.

## Acknowledgments

We thank Mika Shichishima, Yumiko Kito, and Azusa Kamikawa for their contribution to data collection and the staff of the Laboratory Testing Department for their contribution to measuring antibody testing.

