## Supplemental Text 1, Supplemental Figure 1 for "SARS-CoV-2 breakthrough infection during the Delta-dominant epidemic and neutralizing antibodies against Omicron in comparison with the third dose of BNT162b2: a matched analysis"

### Supplementary Appendix

#### Contents

**Supplemental Text 1.** The sources of cells and virus strains........................................................2

**Supplemental Figure 1**. Change in the neutralizing and spike antibody titers against SARS-CoV-2 before and after breakthrough infections stratified by the presence of symptoms............................3

**Supplemental Text 1.** The sources of cells and virus strains

$VeroE6_{TMPRSS2}$ cells were obtained from the Japanese Collection of Research Bioresources (JCRB) Cell Bank (Osaka, Japan) and maintained in Dulbecco's Modified Eagle Medium (DMEM) supplemented with 10% FCS, 100 µg/ml of penicillin, 100 µg/ml of streptomycin, and 1 mg/mL of G418. As the Wuhan strain, the SARS-CoV-2 NCGM-05-2N strain (SARS-CoV-$2^{05\text{-}2N}$) was isolated from nasopharyngeal swabs of a patient with COVID-19 who was admitted to the NCGM hospital in the early phase of the COVID-19 pandemic in 2020. Two clinically isolated SARS-CoV-2 mutant strains, which were provided by the Tokyo Metropolitan Institute of Public Health, Tokyo, Japan, were used in the current study: the B.1.617.2 (Delta) strain [hCoV-19/Japan/TKYK01734/2021 (SARS-CoV-$2^{1734}$, GISAID Accession ID; EPI_ISL_2080609)], the B.1.1.529; BA.1 (Omicron BA.1) strain [hCoV-19/Japan/TKYX00012/2021 (SARS-CoV-$2^{TKYX00012/2021}$, GISAID Accession ID; EPI_ISL_8559478)]. Each variant was confirmed to contain each VOC-specific amino acid substitution before the assays conducted in the present study.

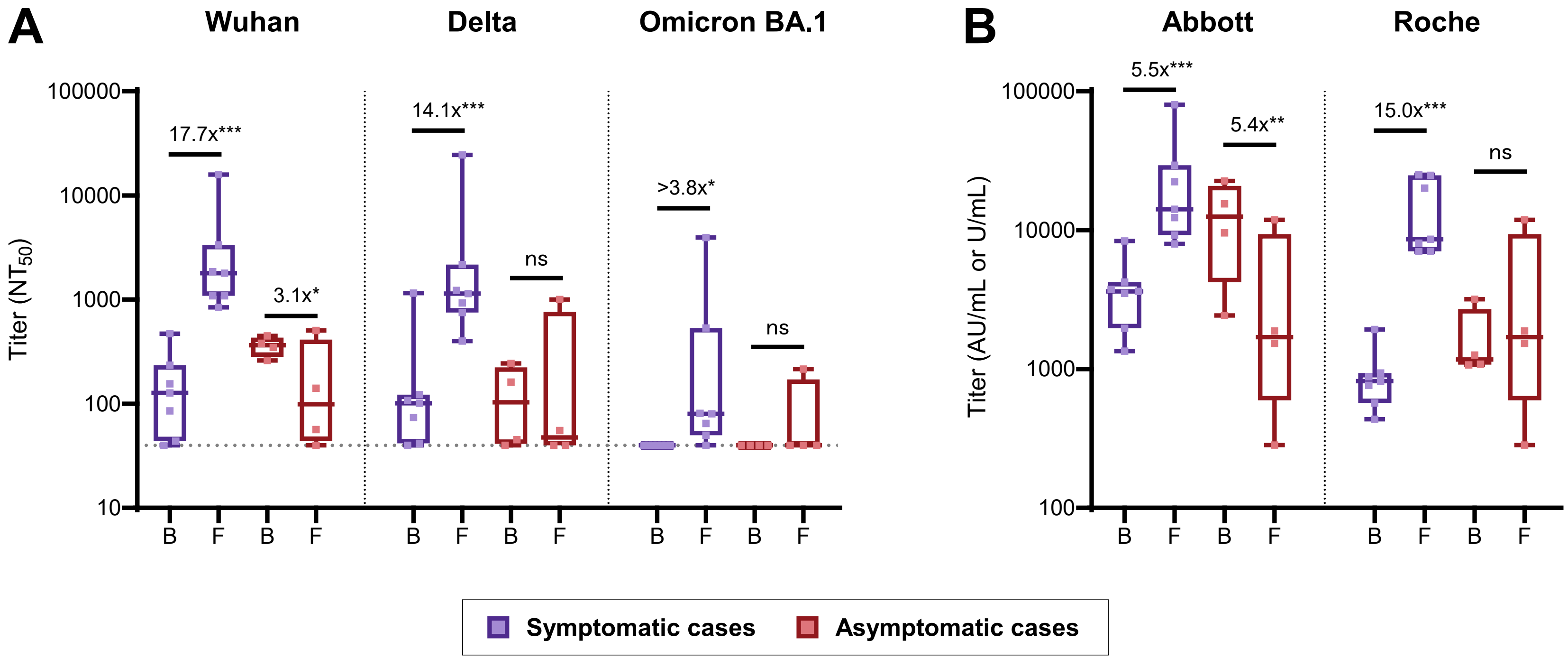


**Supplemental Figure 1**. Change in the neutralizing and spike antibody titers against SARS-CoV-2 before and after breakthrough infections stratified by the presence of symptoms

Panel **A** shows neutralizing antibody titers against the original Wuhan strain, the Delta variant, and the Omicron BA.1 variant determined by 50% focus reduction neutralization test ($FRNT_{50}$) using the serum at baseline and follow-up. Panel **B** shows anti-spike antibody titers measured with the Abbott reagent (AU/mL) and the Roche reagent (U/mL) at baseline and follow-up.

Symptomatic cases correspond to those with positive on PCR testing (n=7), while asymptomatic cases correspond to those with seropositive on any of the anti-SARS-CoV-2 nucleocapsid protein assays (Abbott or Roche assays) at the follow-up survey (n=4).

Box plots show the median, interquartile range, and full range. The dushed horizontal lines indicate the LOD in the present analysis ($NT_{50}$<40 in $FRNT_{50}$).

The fold-change values are estimated ratios of geometric means for antibody titers based on the GEE model (ns: not significant; *P<0.05; **P<0.01; ***P<0.001).

Abbreviations: AU, arbitrary units; B, baseline; F, follow-up; GEE, generalized estimating equation; LOD, limits of detection; $NT_{50}$, 50% neutralization titer; SARS-CoV-2, severe acute respiratory syndrome coronavirus 2.
